# LabARIA: A Multi-Agent System for Integrative Analytics in Clinical Laboratories

**DOI:** 10.64898/2026.11.12.26367970

**Authors:** Haoyang Li, Yaxin Li, Zhenxing Xu, Weishen Pan, Zhen Zhao, Melissa Cushing, Fei Wang, He Sarina Yang

**Author notes:** **Corresponding Author: Fei Wang,** 575 Lexington Ave, New York, NY, 10022, USA, **He Sarina Yang,** 525 E. 68^th^ St, New York, NY, 10065, USA.

## Abstract

**Background:** Laboratory medicine underpins clinical decision-making across nearly every care setting. Clinical laboratory errors and delays can compromise diagnostic reliability and patient care. However, laboratory quality and operational data are commonly reviewed through separate manual workflows, limiting timely recognition of emerging problems across complementary data sources.

**Methods:** We developed and evaluated LabARIA (Laboratory Agentic Reasoning and Integrative Analytics), a multi-agent AI system for coordinated surveillance of specimen workflows, analytical quality control, and laboratory operations. We conducted a single-center retrospective system development and evaluation at NewYork-Presbyterian Hospital/Weill Cornell Medical Center using laboratory data collected from May 1 through June 18, 2026, including 177,480 specimen-panel records, 75,103 internal quality-control (IQC) measurements, 633 proficiency-testing challenges, and 422 recalibration or maintenance actions. Forecasting agents were evaluated using patient-result streams with controlled perturbations, and synthetic staffing and inventory data. We assessed specimen rejection, cancellation, and turnaround-time patterns; warning recall and lead time before documented laboratory actions; assay-specific patient-based real-time quality control (PBRTQC) performance; and exploratory operational forecasting discrimination.

**Findings:** LabARIA characterized specimen and workflow patterns across clinical units and time periods, including hemolysis-related potassium rejections in the intensive care unit during early morning hours (49 of 367 tests [13.3%]). For analytical quality monitoring, the IQC agent generated warnings before 407 of 422 recorded QC actions (recall, 0.964 [95% CI, 0.942-0.978]), with a median lead time of 5.40 days (95% CI, 3.75-6.07). PBRTQC agent selected different monitoring strategies across 35 analytes. In a separate analysis, adding PBRTQC increased action recall from 0.964 to 0.993 (95% CI, 0.979-0.998). A dashboard presented tiered alerts for human review, and an accompanying chatbot supported interactive exploration of the summaries. For the operational forecasting module, areas under the receiver operating characteristic curve for 7-day inventory and staffing risk forecasting were 0.800 (95% CI, 0.696-0.881) and 0.877 (95% CI, 0.778-0.950), respectively.

**Interpretation:** LabARIA demonstrated the feasibility of coordinating laboratory quality surveillance across heterogeneous data sources, providing a foundation for intelligent clinical laboratories. Prospective studies are warranted to determine whether this approach improves monitoring beyond conventional methods, supports laboratory decisions, and benefits patient care without excessive alert burden.

**Funding:** No external funding.

**Research in context:** *Evidence before this study:* We searched PubMed and arXiv for studies published between Jan 1, 2016, and Nov 1, 2026, without language restrictions, using (“clinical laboratory” OR “clinical laboratories” OR “laboratory medicine”) AND (“artificial intelligence” OR “machine learning” OR “large language model” OR “large language models” OR “agentic AI” OR “multi-agent” OR “multiagent” OR “quality control” OR “laboratory operations”). We included studies addressing laboratory quality monitoring or operations and excluded those focused solely on disease diagnosis or treatment selection. Reliable clinical laboratory services are essential to patient care, yet quality and operational surveillance often relies on manual and retrospective review of data distributed across separate systems, placing substantial burdens on laboratory personnel and contributing to reactive responses to emerging problems. Although laboratory automation has advanced, artificial intelligence (AI) and machine learning applications in clinical laboratories have generally focused on narrowly defined tasks, such as identifying preanalytical errors, supporting test interpretation, and improving test utilization. Existing approaches typically rely on predefined rules, descriptive dashboards, or task-specific analytical models rather than coordinated surveillance across laboratory workflows. We identified no published study that had developed and evaluated an integrated agentic AI system for laboratory-wide quality and operational surveillance.

*Added value of this study:* To our knowledge, this is the first study to develop and evaluate a multi-agent AI system for coordinated laboratory-wide quality and operational surveillance. We developed LabARIA to coordinate specialized agents across specimen workflows, analytical quality monitoring, and laboratory operations, integrating their findings into an interactive dashboard with prioritized alerts, supporting measurements, and recommended follow-up checks for laboratory personnel. Rather than providing descriptive summaries alone, the system brings together complementary analytical findings to support the identification and prioritization of emerging concerns. Using real-world clinical laboratory records, we assessed monitoring performance against documented laboratory actions and established quality-control methods, demonstrating early warnings and complementary warning coverage. We also explored operational risk forecasting using synthetic data. Together, these evaluations provide a practical approach for assessing agentic AI in clinical laboratories.

*Implications of all the available evidence:* Our findings suggest that agentic AI could help advance clinical laboratory quality surveillance from fragmented and reactive review toward more coordinated and proactive monitoring. By integrating complementary monitoring functions and presenting prioritized findings with supporting evidence, agentic systems offer a potential approach to helping laboratory personnel identify emerging concerns and focus their investigations. This study advances agentic laboratory surveillance from concept to implementation and evaluation, representing a step toward intelligent clinical laboratories in which AI supports coordinated quality monitoring and human decision-making.

## Introduction

Clinical laboratory testing is essential to disease diagnosis, treatment selection, and monitoring across nearly every care setting.^1^ Errors in specimen collection and handling, analytical quality problems, and delays in test reporting can lead to repeat testing, delay clinical decision-making, prolonged hospital stays, increase resource use, and expose patients to preventable harm.^2,3^ Strengthening laboratory operations is therefore not only a laboratory priority, but an essential component of improving health care delivery. However, maintaining reliable laboratory services has become increasingly challenging because of rising testing volumes, growing diagnostic complexity, and persistent workforce shortages.^4,5^ These challenges underscore the importance of timely identification and investigation of laboratory quality and operational problems.

Clinical laboratories generate extensive information relevant to quality and operational surveillance, including specimen rejection and cancellation records, test turnaround times (TAT), internal quality control (IQC) measurements, patient-result distributions, proficiency-testing results, instrument maintenance records, and staffing and inventory data. However, these data are generally monitored through separate systems and workflows, making it difficult to recognize related abnormalities and requiring laboratory personnel to manually integrate information across multiple sources.^6,7^ Consequently, laboratory personnel may struggle to identify which tests require immediate attention, which clinical units exhibit elevated specimen rejection rates or TAT delays, and which reagent shortages threaten testing continuity.

Integrating these signals could help identify converging problems, such as increasing workload with staffing constraints or deteriorating specimen quality with prolonged TAT, before they compromise patient care. Although artificial intelligence (AI) has been applied to specific laboratory tasks,^8,9^ approaches that coordinate complementary monitoring functions and support integrated interpretation of laboratory-wide quality concerns remain insufficiently explored.

Recent advances in agentic AI offer a promising approach to this systems-level challenge by enabling specialized agents to retrieve relevant data, apply analytical tools, interpret domain-specific signals, and coordinate their findings under human supervision.^10–13^ In this study, we developed and retrospectively evaluated LabARIA (Laboratory Agentic Reasoning and Integrative Analytics), a multi-agent system that brings together patient and specimen monitoring, IQC and patient-based real-time quality control (PBRTQC), proficiency testing and instrument data, and staffing and inventory assessments. Its orchestration layer directs tasks to specialized agents and presents findings with supporting measurements and source context for laboratory personnel to review. We sought to establish the feasibility and initial analytical performance of coordinated laboratory surveillance as a foundation for future prospective evaluation of its incremental clinical and operational value.

## Methods

### Study Design, Setting, Data, and Ethics

We conducted a single-center retrospective development and evaluation of LabARIA at NewYork-Presbyterian Hospital/Weill Cornell Medical Center using laboratory records collected from May 1 through June 18, 2026.^14^ Our study included three components: (1) agent-assisted surveillance and interactive review of specimen quality, test cancellations, and TAT performance using routine laboratory records, with prioritized dashboard alerts and chatbot-supported exploration of findings and supporting evidence; (2) retrospective assessment of analytical QC warnings using historical IQC measurements, patient-result data, and documented laboratory actions; and (3) exploratory operational risk forecasting using synthetic staffing and inventory data. Controlled perturbations were also introduced into patient-result streams to evaluate PBRTQC monitoring. Details are in eMethods 1.

Source data used for building and evaluating LabARIA included laboratory results and specimen records, IQC measurements and documented actions, cancellations, maintenance and recalibration records, proficiency testing records, and local laboratory rules, which comprised 177,480 specimen–panel records from 16 orderable tests and panels spanning 54 analytes; the 8 highest-volume orderables, i.e. basic and comprehensive metabolic panels, complete blood count, magnesium, phosphorus, hepatic function panel, high-sensitivity troponin T, and lactate dehydrogenase, accounted for 92.1% of these records.

This study was approved by the Weill Cornell Medicine Institutional Review Board (21-03023422), with a waiver of informed consent for secondary use of de-identified clinical laboratory data.

### LabARIA System

LabARIA uses a large language model (LLM)–based Orchestrator to coordinate specialized laboratory monitoring and analytical agents (**Figure 1**). <u>The Orchestrator</u> determines the relevant clinical units, laboratory tests, monitoring windows, and data sources; assigns data preparation, laboratory-rule retrieval, analysis, and forecasting tasks to specialized agents, and assembles their findings for laboratory personnel review. <u>The Data Coordinator</u> retrieves laboratory records required for these analyses from the laboratory information system (LIS), instrument middleware, maintenance logs, proficiency testing data, and inventory and staffing records. Specialized agents retrieve and analyze laboratory data using domain-specific rules, statistical monitoring procedures, and forecasting methods. Thus, the agentic architecture provides coordination and integration of analytical workflows, while individual QC signals are generated through their respective analytical procedures. The system is designed to present laboratory findings together with supporting measurements and source context for human interpretation.

**Figure 1.**
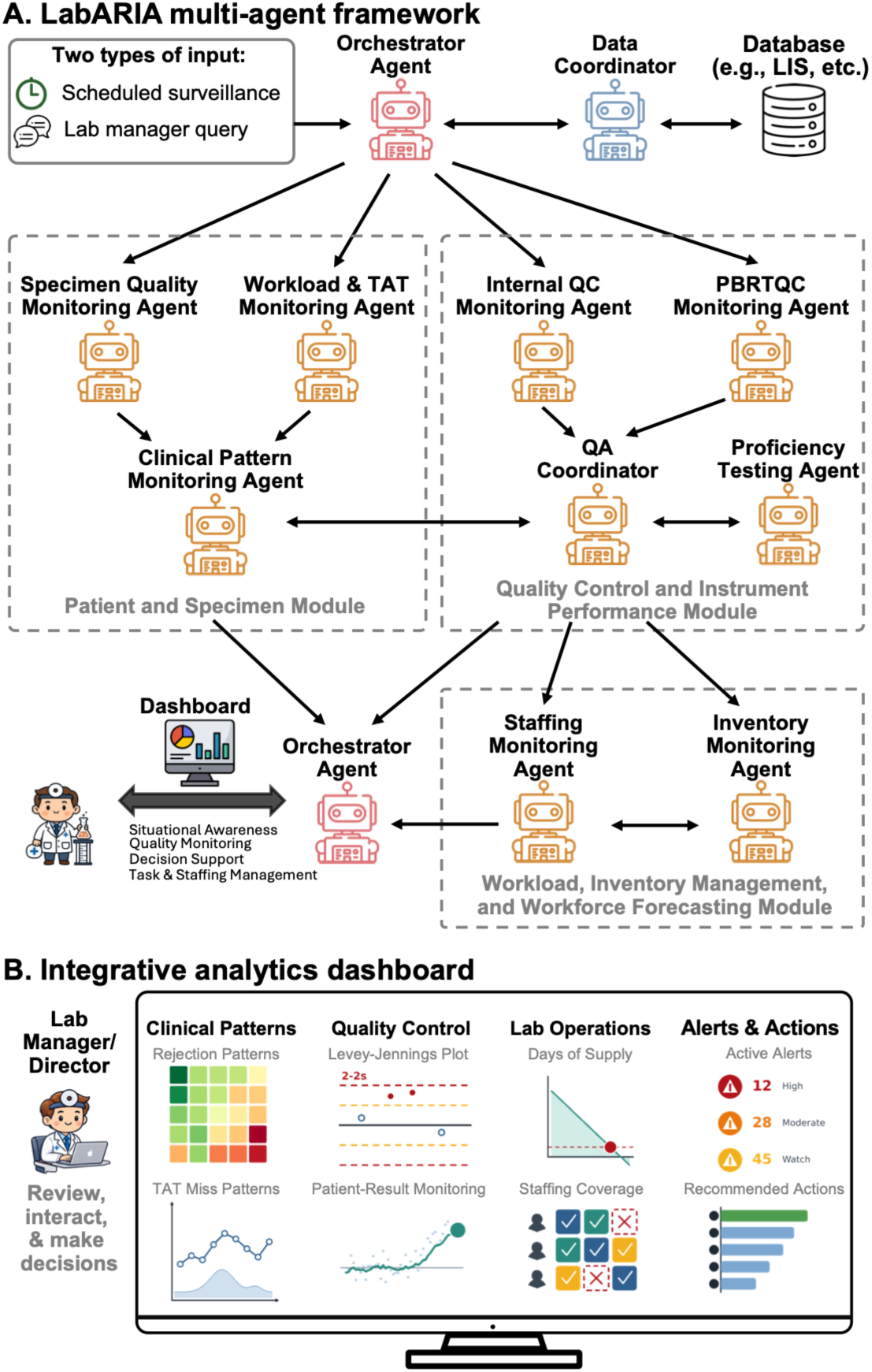
LabARIA Multi-agent System and Dashboard. **A**, Multi-agent system coordinated by the Orchestrator. Scheduled surveillance or a laboratory manager’s query initiates analyses. The Orchestrator works with the Data Coordinator to retrieve relevant laboratory records and assign tasks across 3 functional modules. The Patient and Specimen Module integrates specimen quality, workload, and turnaround time (TAT) findings through the Clinical Pattern Monitoring Agent. The Quality Control and Instrument Performance Module integrates internal quality control (IQC), patient-based real-time quality control (PBRTQC), proficiency-testing results, and instrument records through the QA Coordinator. The Workload, Inventory, and Workforce Module predicts inventory and staffing shortage risks. **B**, Dashboard presenting findings from these modules, supporting measurements, alerts, and recommended checks for laboratory personnel to review.

The LabARIA system consists of three functional modules.^6^ In the <u>Patient and Specimen</u> <u>Module</u>, the Specimen Quality Monitoring Agent tracks preanalytical problems of each clinical unit, including specimen rejections and cancellations attributable to specimen quality or handling, while the Workload & TAT Monitoring Agent independently evaluates testing volume, workload patterns, TAT performance, and delays over time. The Clinical Pattern Monitoring Agent synthesizes their findings to identify affected clinical units, tests, and time periods, retrieve supporting records, and track whether patterns persist, worsen, or resolve.

In the <u>Quality Control and Instrument Performance Module</u>, the QA Coordinator Agent integrates IQC, PBRTQC, proficiency-testing results, and instrument records. The IQC Monitoring Agent evaluates IQC performance and identifies emerging trends across consecutive control results, including sustained shifts, gradual drift, and increasing variability.^15,16^ The PBRTQC Monitoring Agent analyzes patient-result distributions to detect potential analytical drift, systematic bias, and performance instability.^17–19^ The Proficiency Testing Agent reviews proficiency-testing results and incorporates them into longitudinal quality surveillance.^20,21^ These complementary data streams are typically stored in separate files and manually reviewed independently. By integrating QC materials, patient results, and external assessment data across time windows and linking findings to instrument events, the QA coordinator reconciles conflicting signals, estimates confidence, and determines with emerging analytical-quality concerns warrant escalation to laboratory personnel.

In the <u>Workload, Inventory, and Workforce Module</u>, the Staffing and Inventory Monitoring Agents compare anticipated testing demand with shift coverage, staff skills, and available supplies, including reagents, calibrators, and QC materials. In routine practice, staffing schedules, workload data, and inventory records are often managed in separate systems and reviewed independently. This module brings these data together to identify potential mismatches between testing demand and available personnel or supplies.

The Orchestrator consolidates findings from the three modules onto a shared dashboard, including the affected clinical unit or assay, relevant time window, quantitative evidence, comparison thresholds or historical baselines, and recommended follow-up checks (eFigure 1). An accompanying chatbot supports interactive exploration of the summaries. Laboratory personnel retain responsibility for interpreting findings and determining appropriate actions. eFigure 2 shows detailed data sources and agent layers.

**Figure 2.**
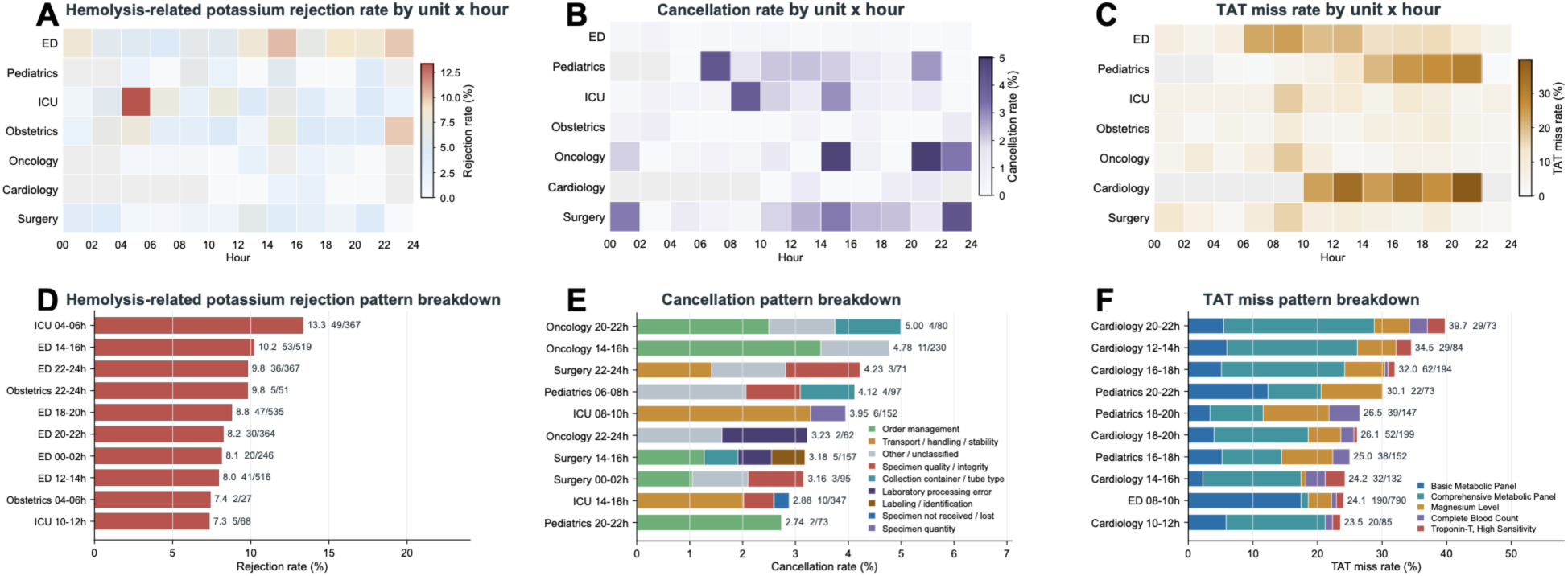
Patient and Specimen Patterns Across Clinical Units and 2-Hour Intervals. **A-C**, Hemolysis-related potassium rejection rates, test cancellation rates, and turnaround time (TAT) miss rates across clinical units and 2-hour intervals during the 28-day window (selected by the user) ending May 31, 2026. **D**, Hemolysis-related potassium rejection rates by clinical unit and time of day. **E**, Cancellation patterns characterized by recorded reasons. **F**, TAT misses characterized by contributing test. ED, emergency department; ICU, intensive care unit.

### Evaluation Designs and Outcomes

We evaluated LabARIA across complementary laboratory surveillance tasks, with outcomes defined according to the available data and reference information.

#### Clinical Pattern Monitoring Across Patient and Specimen Findings

We evaluated the Patient and Specimen Module’s ability to characterize laboratory-recorded quality and workflow patterns across clinical units, assays, and time intervals. Hemolysis-related potassium rejection rates were reported, cancellation findings were characterized by recorded reasons, and TAT miss patterns were broken down by test. The dashboard provided historical comparisons, supporting records, and chatbot-based follow-up (eFigure 1). Details are in eMethods 2.

#### Integrating Quality Monitoring Across IQC, PBRTQC, and Proficiency Testing

The Quality Control and Instrument Performance Module integrated IQC, PBRTQC, proficiency testing, instrument maintenance, and assay recalibration records.

##### IQC Monitoring

Analytical quality monitoring was assessed by evaluating whether the IQC Monitoring Agent could identify emerging QC abnormalities and generate warnings before documented recalibration or maintenance actions. These actions served as reference events for retrospective evaluation. The primary comparator was Westgard rejection rule^22^ routinely used at the study laboratory, defined as either 1 QC result exceeding 3 times standard deviations (SDs) from the target mean (denoted as Westgard 1:3s) or 2 QC results on the same day exceeding the same upper or lower 2-SD limit (Westgard 2:2s). Additional comparators included Westgard 4:1s (4 consecutive results exceeding the same ±1 SD limit in the same direction) and Westgard 10x (10 consecutive results on the same side of the target mean).^22^ Outcomes were action recall, defined as the proportion of QC actions preceded by a warning within the preceding 60 QC results, and warning lead time, defined as the time between warning onset and the recorded QC action. Details are in eMethods 3.

##### Assay-Specific PBRTQC Monitoring

We evaluated the PBRTQC Monitoring Agent by assessing how well it detected shifts in longitudinal patient-result distributions. Following established PBRTQC evaluation approaches,^18,23–25^ we introduced a 5% mean shift in the patient-result distributions to simulate a systematic bias affecting patient results and to assess whether and how quickly the PBRTQC Monitoring Agent detected the bias.

We measured sensitivity, defined as the proportion of simulated shifts detected, the average number of patient results affected before error detection (ANPed^18,23^), calculated as the number of patient results from shift onset to detection, and false-alert frequency before the simulated shift.^26–28^ Details are in eMethods 4.

#### Operational Risk Forecasting

We evaluated whether the Workload, Inventory, and Workforce Module could forecast inventory and staffing risk before disruptions to laboratory testing. The evaluation used synthetic staffing and inventory data covering May 1 through June 18, 2026. Inventory risk was defined as available supply falling below testing needs, and staffing risk was defined as staffing coverage below the level required for the workload. Details are in eMethods 5.

### Statistical Analysis

Performance estimates were reported with 95% confidence intervals (CIs)^29^ using 2000 bootstrap resamples.^30,31^ All statistical tests were 2-sided, with statistical significance indicated as \**P*<.05, \*\**P*<.01, and \*\*\**P*<.001. Analyses were performed using Python, version 3.11.14;^32^ pandas, version 2.3.3;^33^ and NumPy, version 2.4.6.^34^ Details are in eMethods 6.

## Results

During the 28-day monitoring interval ending May 31, 2026, LabARIA characterized specimen rejection, test cancellation, and turnaround-time (TAT) patterns across all laboratory tests and clinical units with flexible monitoring across different time windows (2-hour intervals shown in **Figure 2**). For instance, for hemolysis-related potassium rejections, the highest observed rate occurred in the intensive care unit (ICU) from 4 to 6 AM (49 of 367 tests [13.3%]), followed by the emergency department from 2 to 4 PM (53 of 519 tests [10.2%]) and from 10 PM to midnight (36 of 367 tests [9.8%]). (**Figure 2A, D**)

Cancellation and TAT analyses similarly identified variation across clinical units and time periods. In Oncology, cancellation rates reached 5.0% from 8 to 10 PM (4 of 80 records) and 4.8% from 2 to 4 PM (11 of 230 records), with order-management reasons accounting for the largest component (**Figure 2B, E**). In Cardiology, the highest observed TAT miss rate was 39.7% from 8 to 10 PM (29 of 73 results), predominantly involving comprehensive and basic metabolic panels (**Figure 2C, F**). The system presented these findings alongside historical comparisons and supporting records for laboratory personnel to review.

The IQC Monitoring Agent identified patterns of sustained deviation, progressive drift, and increasing variability in longitudinal control measurements (**Figure 3A-C**). Among 422 documented recalibration or maintenance actions, 407 (96.4% [95% CI, 94.2%-97.8%]) were preceded by a warning within the preceding 60 QC results. The median interval between warning onset and recorded action was 5.40 days (95% CI, 3.75-6.07 days). For comparison, the reported median lead times were 0.037 days for the laboratory’s routine Westgard 1:3s or 2:2s rule, 0.481 days for Westgard 4:1s, and 4.071 days for Westgard 10x (**Figure 3D-E**).

**Figure 3.**
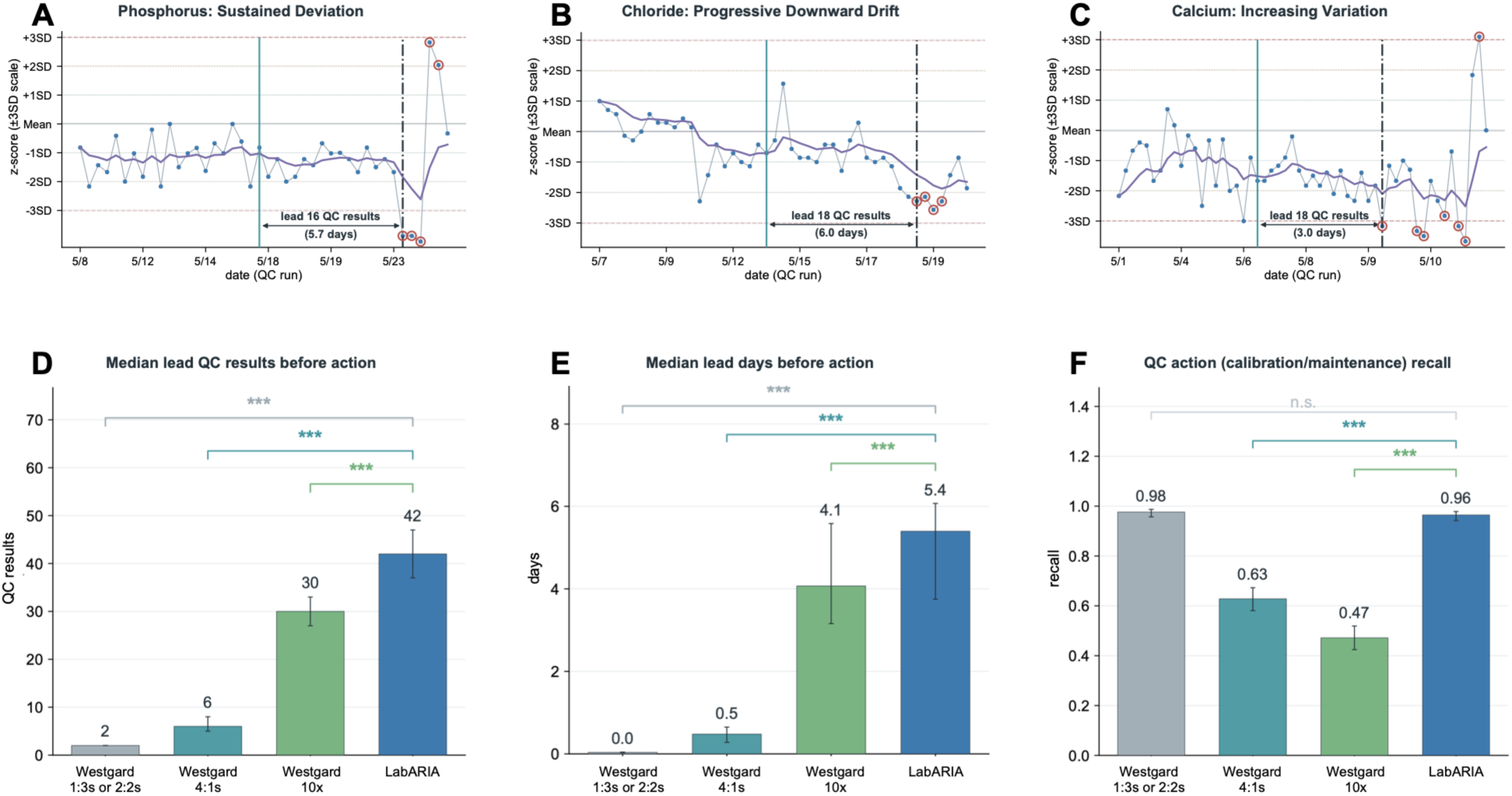
Internal Quality Control Warning Recall and Lead Time Before Recorded QC Actions. A-C, Phosphorus, chloride, and calcium examples illustrating three patterns in consecutive control results identified by the Internal QC Monitoring Agent: a sustained deviation from the target mean (A), a progressive upward or downward drift (B), and increasing variation (C). Blue points indicate QC results, purple lines indicate exponentially weighted moving average (EWMA) traces, and open red circles indicate QC results meeting the Westgard rejection rule (Westgard 1:3s or 2:2s). Green vertical lines indicate warning onset from the Internal QC Monitoring Agent, and black dashed vertical lines indicate recorded QC actions (recalibration or maintenance). **D-E**, Median warning lead time before recorded QC actions, measured in QC results (D) and days (E), comparing the Internal QC Monitoring Agent with the Westgard 1:3s or 2:2s, Westgard 4:1s, and Westgard 10x. F, QC action recall, defined as the proportion of recorded QC actions preceded by a warning within the preceding 60 QC results. Error bars indicate 95% confidence intervals (CIs). SD, standard deviation. \**P*<.05, \*\**P*<.01, and \*\*\**P*<.001; n.s., not significant.

These results indicate the monitoring procedure generated early signals associated with subsequent documented laboratory actions.

Patient-result distributions differed substantially across the 35 analytes evaluated, with coefficients of variation^35,36^ ranging from 2.8% for sodium to 85.8% for troponin T (**Figure 4A**).

**Figure 4.**
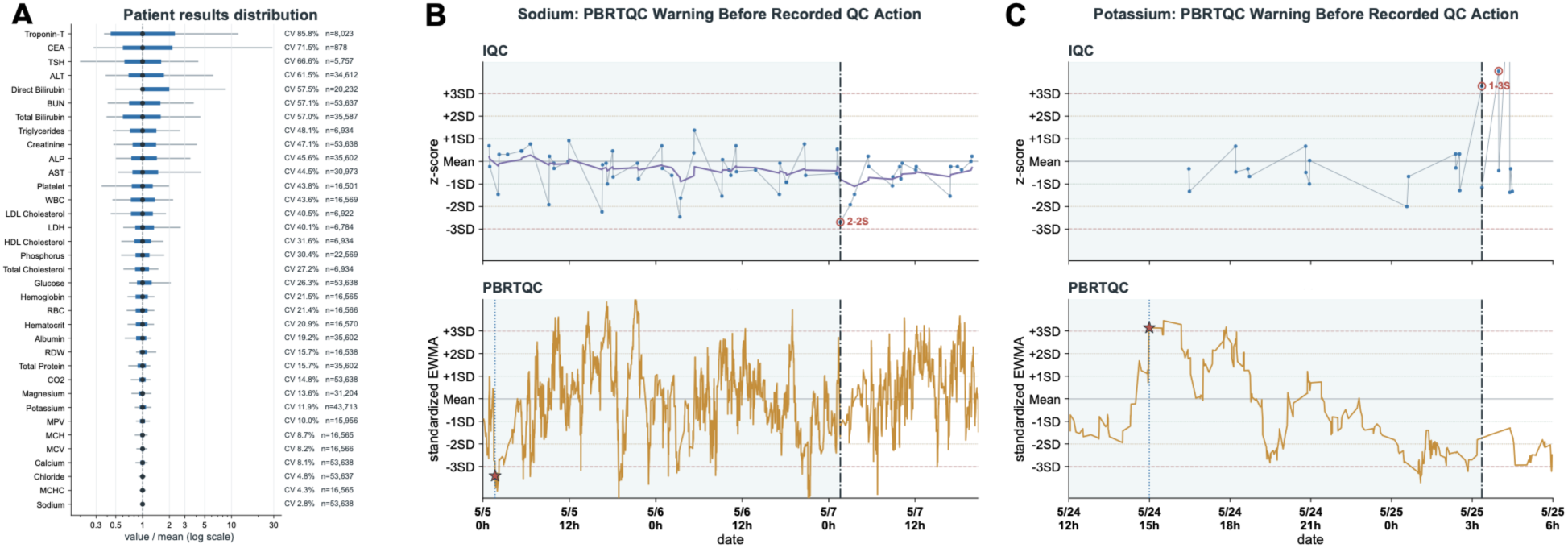
Patient-Result Distributions and Complementary PBRTQC Warnings. **A**, Patient-result distribution for each analyte. Points indicate medians, dark intervals indicate the 25th to 75th percentiles, and light lines indicate the 5th to 95th percentiles. Patient results were normalized to each analyte’s mean and displayed on a logarithmic scale. Coefficients of variation (CVs) were calculated as the ratio of the standard deviation to the mean and reported as percentages; n indicates the number of patient results. **B-C**, Sodium (B) and potassium (C) examples illustrating patient-based real-time quality control (PBRTQC) warnings before recorded QC actions. Upper panels show internal quality control (IQC) results, and lower panels show PBRTQC monitoring. Blue points indicate QC results, the purple line indicates an exponentially weighted moving average (EWMA) trace, and open red circles indicate QC results meeting the Westgard rejection rule (Westgard 1:3s or 2:2s). Orange lines indicate PBRTQC EWMA traces, and stars indicate PBRTQC warnings. Blue dotted vertical lines indicate PBRTQC warning onset, and black dashed vertical lines indicate recorded QC actions (recalibration or maintenance). For sodium, PBRTQC generated a warning approximately 2 days before the recorded action when no warning was generated by the Internal QC Monitoring Agent. For potassium, PBRTQC generated a warning approximately 0.5 days before the recorded action. SD, standard deviation.

These differences motivated assay-specific selection of PBRTQC monitoring strategies.^37^ In representative sodium and potassium examples, PBRTQC generated warnings before documented laboratory actions when IQC monitoring had not generated a qualifying warning (**Figure 4B-C**).

In controlled perturbation experiments involving a 5% shift in patient-result means, PBRTQC achieved a detection sensitivity of 67.5% ± 16.8% across 35 analytes, with a mean ANPed of 72.6 ± 25.7 patient results and a false-alert frequency of 0.44 ± 0.37 per 1000 patient results before the shift. Performance varied across analytes, with sodium showing the highest sensitivity (100%; ANPed, 8.0) and lactate dehydrogenase the lowest sensitivity (45%; ANPed, 99.8).

Across all documented QC actions, PBRTQC generated additional warnings before 12 of the 15 actions not preceded by an IQC warning. Combining PBRTQC with IQC increased the proportion of documented actions preceded by warnings from 407 of 422 (96.4%) to 419 of 422 (99.3% [95% CI, 97.9%-99.8%]). These findings demonstrate complementary warning coverage across the two monitoring approaches, although the effect of combining them on overall false-alert frequency was not established.

Using synthetic staffing and inventory data, the Workload, Inventory, and Workforce Module estimated the risk of inventory depletion and insufficient staffing coverage over 3-, 7-, and 14-day horizons. For the 7-day prediction horizon, the areas under the receiver operating characteristic curve^38,39^ were 0.800 (95% CI, 0.696-0.881) for inventory risk and 0.877 (95% CI, 0.778-0.950) for staffing risk. Discrimination decreased at longer prediction horizons (eFigure 4).

## Discussion

In this single-center retrospective study, we developed and evaluated LabARIA, a multi-agent AI system designed to coordinate surveillance across clinical laboratory quality and operational workflows. Using routine laboratory records, the system characterized specimen rejection, cancellation, and TAT patterns across clinical units; generated warnings preceding most documented recalibration or maintenance actions; and identified additional actions through complementary patient-based quality-control monitoring. Exploratory analyses also demonstrated the feasibility of forecasting operational risks using synthetic staffing and inventory data. Together, these findings establish initial technical feasibility and characterize the analytical performance of several monitoring components within an integrated laboratory surveillance framework.

The principal motivation for LabARIA is the fragmentation of laboratory quality surveillance. Laboratory personnel must routinely assess specimen quality, analytical performance, instrument maintenance, testing demand, and operational resources, frequently using information distributed across separate systems. Although individual monitoring methods can identify abnormalities within specific processes, laboratory investigation may require integration of several complementary sources of evidence. For example, increasing specimen rejection rates without corresponding abnormalities in analytical QC measurements may suggest a preanalytical problem, whereas concurrent changes in patient-result distributions and internal quality control may raise concern about analytical performance. By coordinating specialized monitoring tasks and presenting their findings within a shared laboratory context, LabARIA offers an approach to supporting integrated assessment of such problems (**Figure 5**). The two illustrative scenarios in eAppendix 1 in Supplement 1 demonstrate how complementary evidence from specimen quality, analytical monitoring, and laboratory operations could be synthesized to trouble-shoot an increased hemolysis-related potassium rejection in ED and a sudden change of calcium assay performance, respectively. Although these examples highlight the potential value of cross-module coordination, its incremental benefit over independent monitoring approaches remains to be established empirically.

**Figure 5.**
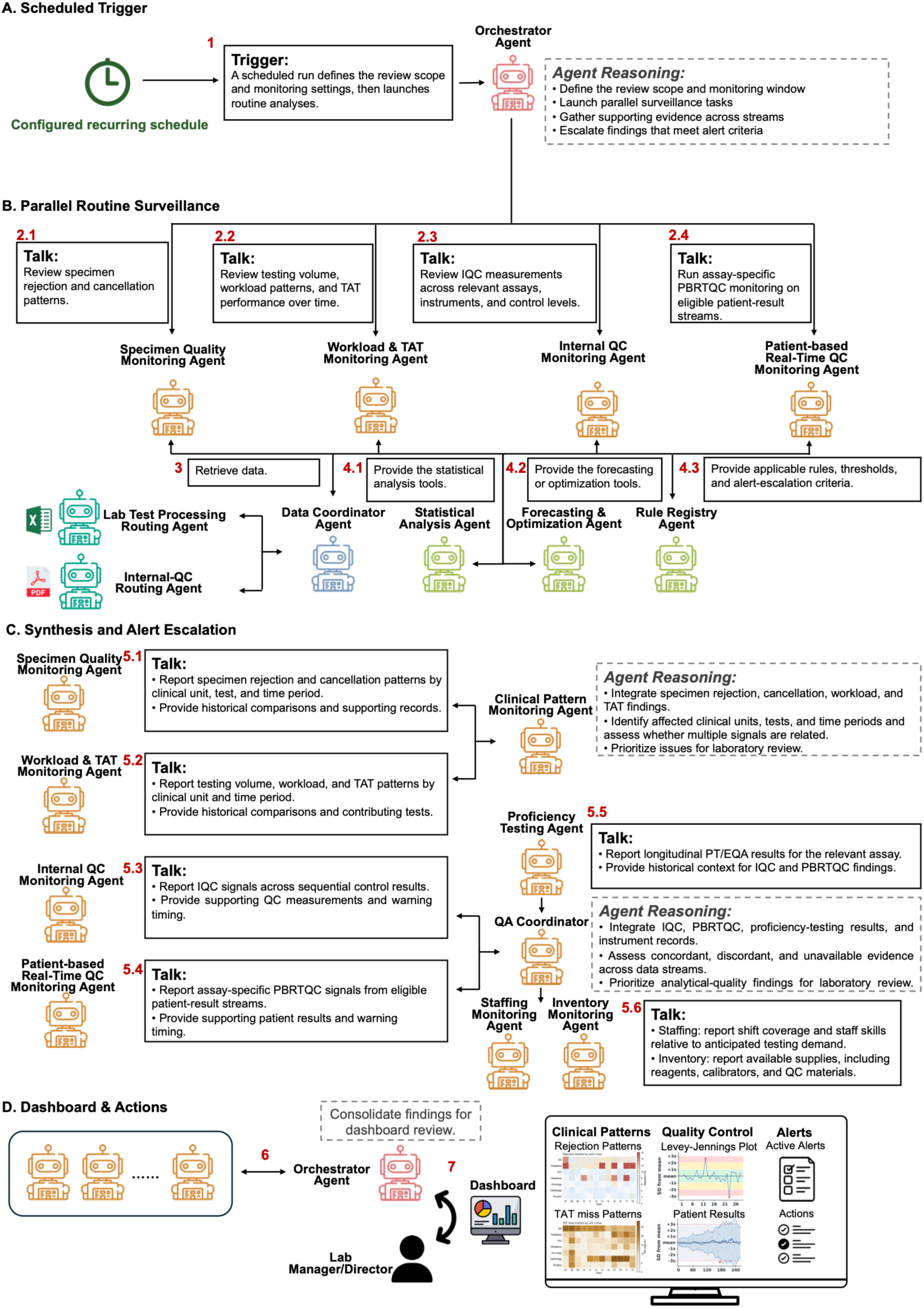
Scheduled-Surveillance Workflow. **A**, Scheduled surveillance initiates the Orchestrator, which determines the review scope, monitoring window, and relevant data sources. **B**, The Orchestrator coordinates parallel monitoring of specimen quality, testing volume and turnaround time (TAT), internal quality control (IQC), and patient-based real-time quality control (PBRTQC). The Data Coordinator retrieves relevant laboratory records, and specialized analytical tools support these tasks. **C**, The Clinical Pattern Monitoring Agent synthesizes specimen quality and workload/TAT findings to identify affected clinical units, tests, and time periods. The QA Coordinator integrates IQC, PBRTQC, proficiency-testing results, and instrument records. The Staffing and Inventory Monitoring Agents compare anticipated testing demand with shift coverage, staff skills, and available supplies. **D**, The Orchestrator assembles findings and supporting measurements for dashboard review.

Current middleware, autoverification system, dashboards, and ML models improve laboratory operation through predefined rules or narrowly specific tasks, whereas agentic AI systems have the capability of dynamically retrieving information, using tools, coordinating across data streams, and adapting actions to evidence and uncertainty under human oversight. The present results also illustrate the potential importance of complementary monitoring methods. IQC warnings preceded 96.4% of documented recalibration or maintenance actions, and the addition of PBRTQC increased combined action coverage to 99.3%. Because IQC and PBRTQC monitor different sources of information, their combination may provide a broader view of analytical quality than either approach alone. However, these findings require cautious interpretation. Documented recalibration or maintenance actions are imperfect reference events for analytical failure, and retrospective action coverage does not establish the diagnostic accuracy of warnings for clinically meaningful quality problems. Moreover, earlier warnings are beneficial only if they support appropriate investigations without generating an excessive burden of false alerts. Prospective evaluation should therefore assess actionable-event detection, false-alert frequency, alert prioritization, and the consequences of laboratory personnel responding to system-generated recommendations.

An important distinction is that the current evaluation establishes the performance of individual monitoring functions but does not directly quantify the incremental benefit of multiagent coordination. The broader value of the architecture depends on whether synthesizing evidence across specimen, analytical, and operational workflows improves the accuracy, efficiency, or prioritization of laboratory investigations compared with conventional monitoring or independently operating analytical tools. Evaluating this added value will require assessments of integrated clinical laboratory episodes, comparisons against existing practices, and measurement of human performance with and without AI assistance. Such studies should examine whether integrated surveillance enables laboratory personnel to identify relevant problems more accurately, investigate them more efficiently, and respond without unnecessary escalation or automation bias.

This study has several limitations. First, it was conducted at a single academic medical center using a relatively short period of historical laboratory data, limiting its generalizability across institutions, instruments, assays, and operating conditions. Second, documented QC actions were used as reference events but were not independently adjudicated as true analytical failures. Consequently, sensitivity, positive predictive value, and false-alert burden for clinically meaningful quality problems could not be established. Third, descriptive specimen and workflow analyses characterized observed patterns but did not evaluate the accuracy of root-cause identification or the clinical importance of detected variation. Fourth, operational risk forecasting was assessed using synthetic staffing and inventory data. Fifth, the accuracy of cross-agent evidence synthesis, effects on laboratory personnel’s decision-making, and downstream clinical and operational outcomes were not directly evaluated. Further studies are needed to assess performance under prospective conditions, characterize failure modes, and establish the reliability of integrated agentic surveillance across different laboratory settings.

In conclusion, LabARIA demonstrates the feasibility of integrating multiple laboratory monitoring functions within a coordinated multiagent AI framework and serving as a cohesive intelligence layer capable of managing the complexity of modern diagnostic operations. Retrospective findings showed early warnings associated with documented quality-control actions and complementary coverage from patient-based monitoring. The modular architecture preserves existing laboratory infrastructure while enabling adaptive coordination, transparent provenance, and incremental deployment. Whether this approach improves laboratory quality, operational efficiency, or patient safety beyond existing surveillance practices remains to be determined through rigorous comparative and prospective evaluation.

## Article Information

### Author Contributions

F.W. and H.Y. conceived the initial idea. H.L. conceived the method and designed the algorithmic techniques. H.L. developed the code and performed the computational analysis. H.L. drafted the initial manuscript; Y.L. provided retrospective laboratory data; F.W. and H.Y. provided critical revisions; Z.Z. and M.C. edited the manuscript; W.P. and Z.X. reviewed the manuscript and provided suggestions. F.W. and H.Y. supervised the project.

All authors reviewed, provided feedback, and approved the final manuscript

### Data Sharing Statement

This study analyzed de-identified clinical laboratory data from Weill Cornell Medicine. Owing to patient privacy and institutional regulations, these data are not publicly available. Access may be provided to qualified researchers upon reasonable request to the corresponding author and subject to approval by the Weill Cornell Medicine Institutional Review Board.

## Acknowledgment

Drs Li and Yang had full access to all the data in the study and take responsibility for the integrity of the data and the accuracy of the data analysis.

