## Supplement 1 for "LabARIA: A Multi-Agent System for Integrative Analytics in Clinical Laboratories"

**LabARIA: A Multi-Agent System for Integrative Analytics in Clinical Laboratories**
**Supplemental Methods, Results, Tables, and Figures**
**Contents**


- **eMethods**
  - eMethods 1. LabARIA System
  - eMethods 2. Clinical Pattern Monitoring Across Patient and Specimen Findings
  - eMethods 3. IQC Monitoring
  - eMethods 4. Assay-Specific PBRTQC Monitoring and Complementary Warning Coverage
  - eMethods 5. Operational Risk Forecasting
  - eMethods 6. Statistical Analysis
- **eResults**
  - eResults 1. Quality Control and Instrument Performance
  - eResults 2. Inventory and Staffing Forecasting
- **eAppendix**
  - eAppendix 1. Illustrative Scenarios of Integrated Laboratory Investigation
- **eTables**
  - eTable 1. IQC Action Recall and Warning Lead Time
  - eTable 2. Sensitivity of IQC Action Recall and Warning Lead Time to Monitoring Thresholds
  - eTable 3. Analyte-Specific PBRTQC Methods and Performance Under a 5% Mean Shift
  - eTable 4. IQC and PBRTQC Action Recall
  - eTable 5. Inventory and Staffing Forecasting Performance
  - eTable 6. Analyte-Specific Hemolysis-related Rejection Cutoffs
  - eTable 7. Test-Specific Turnaround-Time Targets
- **eFigures**
  - eFigure 1. LabARIA Dashboard for Laboratory Personnel
  - eFigure 2. Laboratory Data Sources and Agent Architecture
  - eFigure 3. PBRTQC Monitoring Performance and Complementary Warnings
  - eFigure 4. Inventory and Staffing Monitoring and Forecasting
- **Supplementary References**

### eMethods

#### eMethods 1. LabARIA System

LabARIA uses a large language model (LLM[1,2])–based Orchestrator to coordinate monitoring initiated by scheduled surveillance or a laboratory manager's request. The system retrieves records from the relevant laboratory data sources, applies laboratory rules[3,4] and analytical methods, and presents findings from the three functional modules with supporting measurements and source records for laboratory personnel to review. To protect laboratory data privacy, the implementation used locally served Gemma 4[2] for task coordination, execution, and reasoning, while GPT-5.5[5] supported chatbot-based follow-up using only summarized analytical results (eFigure 1).

##### Agent Roles and Coordination

The agents in eFigure 2 are organized into routing, foundation, and application agents.

Routing agents prepare records for the relevant analytical tasks. The Data Coordinator Agent identifies required data sources, checks their availability for the selected monitoring window, and reports missing records. The Lab Test Processing Routing Agent prepares specimen records by analyte, clinical unit, and time for specimen quality, cancellation, turnaround time (TAT), and patient-based real-time quality control (PBRTQC) analyses. The Internal-QC Routing Agent organizes internal quality control (IQC) measurements and documented quality control (QC) actions by assay, instrument, control material, lot, and time. The Proficiency-Testing Routing Agent organizes proficiency-testing/external quality-assessment (PT/EQA) challenges with their official grades and annotations.

Foundation agents provide analytical and reasoning capabilities used by the application agents. The Statistical Analysis Agent supports summaries, QC trend analysis, warning lead-time assessment, and uncertainty estimates. The Forecasting & Optimization Agent supports inventory and staffing risk forecasting across different time windows. The Rule Registry Agent retrieves applicable laboratory rules and test-specific thresholds, including analyte-specific hemolysis-related rejection cutoffs, TAT targets, and QC rules. The Reasoning & Explanation Agent synthesizes findings across analytical tasks, links them to supporting measurements and source records, and identifies missing or conflicting evidence. It also provides contextual explanations of the findings and supports chatbot-based follow-up questions using summarized analytical results.

Application agents perform laboratory monitoring and integrate analytical findings. The Orchestrator Agent determines the relevant clinical units, tests, monitoring windows, and data sources, assigns tasks to specialized agents, and assembles findings with supporting evidence for laboratory personnel to review. The Specimen Quality Monitoring Agent monitors hemolysis-related specimen rejections based on analyte-specific cutoffs and examines recorded test cancellations and their reasons. The Workload & TAT Monitoring Agent evaluates testing volume, workload patterns, and TAT performance against test-specific targets. The Clinical Pattern Monitoring Agent integrates specimen rejection, cancellation, and TAT findings to identify affected clinical units, tests, and time periods and characterize contributing cancellation reasons and tests associated with TAT misses. The Internal QC Monitoring Agent examines consecutive control results to identify sustained shifts, gradual drift, and increasing variability, linking warnings to recorded QC actions. The PBRTQC Monitoring Agent analyzes patient-result distributions using assay-specific monitoring methods to detect potential analytical shifts and provide warnings complementary to IQC. The Proficiency Testing Agent reviews PT/EQA results and incorporates external assessment findings into longitudinal quality surveillance. The QA Coordinator integrates IQC, PBRTQC, PT/EQA, maintenance, and recalibration findings to assess emerging analytical quality concerns and present recommended follow-up checks. The Staffing Monitoring Agent compares anticipated testing demand with staffing coverage and available staff skills to identify potential staffing shortages. The Inventory Monitoring Agent evaluates available supplies, expected consumption, delivery lead times, and expiration dates to identify potential inventory shortages.

##### Patient and Specimen Module

The Patient and Specimen Module evaluates specimen rejection, test cancellations, testing volume, and turnaround time (TAT) across clinical units and time intervals. Rejection, cancellation, and TAT miss rates are calculated using the corresponding eligible records (eTables 6 and 7). The Clinical Pattern Monitoring Agent compares these rates with laboratory targets and historical performance from the same clinical unit and metric before the selected monitoring window. It also retrieves recorded cancellation reasons and tests contributing to TAT misses. These findings help laboratory personnel distinguish the affected clinical units, tests, and time periods and review whether patterns persist or change as additional records become available.

**Quality Control and Instrument Performance Module**
For IQC, consecutive control results are examined using exponentially weighted moving average (EWMA),[6] cumulative sum (CUSUM),[7–9] and rolling standard deviation (SD)[10] or coefficient of variation (CV)[11] monitoring to assess sustained shifts, gradual drift, and increasing variability. Warnings are linked to affected assays, control measurements, and recorded QC actions and are displayed alongside the laboratory's Westgard rules.[3]
For PBRTQC, the system examines patient-result distributions using rolling mean, rolling median, EWMA, CUSUM, and Bull's trimmed mean.[12–14] Methods are selected separately for each analyte based on the average number of patient results from shift onset to detection (ANPed[12,13]), subject to no more than 3 false alerts per 1000 patient results before the simulated shift, following established PBRTQC evaluation approaches.[12,13] The QA Coordinator considers IQC, PBRTQC, PT/EQA, maintenance, and recalibration findings together, including evidence that agrees, conflicts, or is unavailable, before presenting recommended checks for laboratory personnel.
**Workload, Inventory, and Workforce Module**
The module compares anticipated testing demand with staffing coverage and available staff skills across laboratory sections and shifts, as well as available supplies, including reagents, calibrators, and QC materials. Inventory monitoring considers expected consumption, delivery lead times, and expiration dates. Forecasting is performed over 3-, 7-, and 14-day time windows using simple moving average (SMA), a first-order autoregressive model (AR(1)), and linear trend extrapolation.[15] Forecasted risks are presented with the corresponding workload and available resources.
**Dashboard and Laboratory Personnel Review**
The dashboard presents findings from the three modules as priority-tiered alerts with supporting measurements, laboratory targets or historical baselines, and source records. It supports review by clinical unit, test, assay, and monitoring window (eFigure 1). Laboratory personnel can ask follow-up questions using the displayed summaries and associated analytical results. Interpretation and subsequent actions remain the responsibility of laboratory personnel.
**Outcome Definitions and Evaluation Periods**
Hemolysis-related specimen rejection was assessed using the analyte-specific cutoffs in eTable 6. Cancellations were laboratory-recorded events with order-related or specimen-related reasons, including specimen instability resulting from handling issues or transportation delays, incorrect test ordering, insufficient specimen volume, use of an incorrect collection tube, or compromised specimen integrity. A TAT miss was defined as a test result verified after its predefined test-specific target. Patient and specimen analysis were evaluated within a 28-day window (selected by the user) ending May 31, 2026. The IQC and PBRTQC analyses used laboratory records from May 2026. The warning-coverage analysis included 422 recorded QC actions, defined as recalibration or maintenance. Patient-result streams for the same analyte and instrument were available for 200 of these actions.

**eMethods 2. Clinical Pattern Monitoring Across Patient and Specimen Findings**
The analysis presented in Figure 2 included 7 clinical units: emergency department (ED), pediatrics, intensive care unit (ICU), obstetrics, oncology, cardiology, and surgery. The 28-day monitoring window extended from May 4 through May 31, 2026. For each clinical unit, records were grouped into 12 consecutive 2-hour intervals according to time of day. Hemolysis-related rejection rates were calculated as the number of rejected tests divided by all tests within each clinical unit and interval. Analyte-specific hemolysis-related rejection cutoffs are listed in eTable 6. For example, the potassium hemolysis rejection cutoff was an H-index greater than 100 mg/dL, meaning that potassium test results were rejected when the specimen's H-index exceeded this threshold. Cancellation rates were calculated as the number of recorded cancellations divided by all received specimen-test records. TAT miss rates were calculated as the number of results exceeding the test-specific target divided by all test-result records with applicable TAT targets (eTable 7).

**eMethods 3. IQC Monitoring**
The default monitoring settings included an EWMA[6] limit of 1.0, a CUSUM[7–9] decision interval of 4.0, and a rolling standard deviation (SD) or coefficient of variation (CV) limit of 1.5. Action recall was defined as the proportion of recorded QC actions preceded by a warning within the previous 60 QC results. A warning episode began when a monitoring threshold was exceeded and ended when the signal cleared. Warning lead time was measured from the onset of the earliest warning to the recorded QC action,

expressed in both QC results and days (eTable 1). Warnings without a corresponding recorded action were not used to estimate IQC false-alert frequency.

To assess sensitivity to monitoring settings, the EWMA limit (0.75, 1.0, and 1.25), CUSUM decision interval (3.0, 4.0, and 5.0), and rolling SD[10] or CV[11] limit (1.25, 1.5, and 1.75) were varied individually while the remaining settings were held at their default values (eTable 2).

**eMethods 4. Assay-Specific PBRTQC Monitoring and Complementary Warning Coverage**

We evaluated PBRTQC detection performance by introducing a simulated 5% mean shift into recorded patient-result streams from 35 analytes, following established PBRTQC evaluation approaches.[12,13] Each evaluation included 360 patient results, with the shift introduced after the first 221 results and the remaining 139 results used to assess detection. Analyte-specific reference values were estimated from recorded results and held constant across repeated evaluations. A simulated shift was considered detected when the monitoring statistic first crossed the applicable limit after shift onset.

False-alert frequency was expressed as alerts per 1000 patient results before the simulated shift. For each analyte, the PBRTQC Monitoring Agent selected the method with the lowest ANPed among those producing no more than 3 false alerts per 1000 patient results before the shift. The selected methods and their performance are reported in eTable 3 and eFigure 3A.

For Figure 4A, patient-result distributions were normalized to each analyte's mean and displayed on a logarithmic scale. CV was calculated as the ratio of SD to the mean and reported as a percentage.

We evaluated whether PBRTQC identified additional recorded QC actions beyond IQC monitoring using the same 422 recorded QC actions. We compared action recall for IQC alone and combined IQC and PBRTQC monitoring. For combined monitoring, an action was counted as identified when either IQC or PBRTQC generated a warning before the recorded action.

**eMethods 5. Operational Risk Forecasting**

The operational evaluation used 49 days of synthetic inventory and staffing data from May 1 through June 18, 2026. Inventory data included 300 catalog items, and staffing data represented 2 laboratory sections and 3 shifts. For inventory monitoring, severity was categorized as low when days of supply was below the reorder threshold but remained at or above the safety-stock threshold, moderate when it was below the safety-stock threshold but remained at or above the delivery lead time, high when it was below the delivery lead time but above zero, and critical when stock reached zero or an expired lot remained in use.

Forecasting was evaluated at 3-, 7-, and 14-day time windows. For inventory risk forecasting, an inventory shortage was defined as available supply falling below testing needs. The evaluated outcome was whether an inventory shortage occurred within the time window. For staffing risk forecasting, the evaluated outcome was whether staffing coverage fell below the level required for the workload within the same time window. Each inventory record represented one inventory item on a given day, and each staffing record represented one laboratory section and shift on a given day.

The Forecasting & Optimization Agent evaluated 3 candidate forecasting methods for each outcome and time window: simple moving average (SMA), a first-order autoregressive model (AR(1)), and linear trend extrapolation. For calculations using recent observations, the most recent 7 days, including the prediction day, were used; when fewer than 7 days were available, all available observations were used.

For inventory risk forecasting, SMA estimated daily consumption as the mean consumption over the most recent 7 days. Days of supply was calculated by dividing current stock by the estimated daily consumption, and the negative days-of-supply value was used as the risk score. The AR(1) model forecast daily consumption using all observations available through the prediction day. The historical mean and lag-1 Pearson correlation were estimated from the available series and used recursively to forecast future consumption. Negative forecast consumption was set to zero, cumulative forecast consumption was subtracted from current stock, and the risk score was defined as the negative of the minimum predicted stock within the specified time window. Linear trend extrapolation estimated a least-squares slope from recent stock levels and calculated the value at the end of the time window as the current stock plus the estimated slope multiplied by the length of the time window. The negative of the predicted stock at the end of the time window was used as the risk score.

For staffing risk forecasting, SMA calculated mean staffing coverage over the most recent 7 days, and the negative mean coverage was used as the risk score. The AR(1) model recursively forecast staffing coverage using all observations available through the prediction day, and the risk score was defined as the negative of the minimum predicted staffing coverage within the specified time window. Linear trend extrapolation estimated a least-squares slope from recent staffing coverage and calculated coverage at the end of the time window as the current coverage plus the estimated slope multiplied by the length of the time window. The negative of the predicted staffing coverage at the end of the time window was used as the risk score.

For each outcome and time window, forecasting-method selection was based on AUROC calculated from observations whose outcome windows had closed by the final prediction day.

We compared the selected forecasting method with 3 baseline approaches that assigned risk scores to each inventory or staffing record. The random baseline generated uniformly distributed random scores. The time-based baseline assigned higher scores with increasing time since the last restocking or the last day with sufficient staffing. The historical-frequency baseline calculated risk scores using the proportion of prior days with inventory or staffing problems for the corresponding inventory item or laboratory section and shift. For each time window, forecasting performance was evaluated by comparing the predicted risk rankings with whether inventory or staffing shortages occurred within the corresponding future time window.

Forecasting performance was evaluated primarily using the area under the receiver operating characteristic curve (AUROC). Additional measures were average precision (AP), Recall@K, Spearman rank correlation ($\rho$), Kendall rank correlation ($\tau$), and normalized discounted cumulative gain (nDCG).[16–19] AP summarizes precision across recall levels. Recall@K measures the proportion of records with future inventory or staffing shortages identified among the K highest-ranked records, where K equals the number of records with these outcomes. nDCG gives greater weight to records with future inventory or staffing shortages ranked nearer the top. Spearman and Kendall correlations assess the association between predicted risk scores and observed outcomes. These measures complement AUROC by evaluating how well the methods rank future inventory and staffing risks (eFigure 4).

**eMethods 6. Statistical Analysis**

Patient and specimen monitoring results were summarized by clinical unit and time interval. QC action recall was reported with Wilson 95% confidence intervals (CIs), which account for the bounded range of proportions and allow asymmetric confidence limits, particularly for proportions close to 0 or 1.[20] For warning lead times and differences in warning lead time, CIs were calculated using the 2.5th and 97.5th percentiles of bootstrap estimates. This method allows confidence limits to reflect the distribution of bootstrap estimates without assuming a normal distribution.[21,22] Bootstrap resampling was performed at the recorded QC action level, and comparisons of warning lead time included only actions identified by both methods. Exact McNemar tests were used to compare action recall between IQC alone and combined IQC and PBRTQC monitoring, and between IQC and PBRTQC monitoring. These tests were used because each recorded QC action was evaluated using the two monitoring approaches being compared, and the number of actions identified by only one approach was small.[23,24]

### eResults

#### eResults 1. Quality Control and Instrument Performance

##### IQC Monitoring Warned Before Recorded QC Actions

Across the 422 recorded QC actions, action recall was 0.976 (95% CI, 0.957–0.987) for Westgard 1:3s or 2:2s, 0.628 (95% CI, 0.581–0.673) for Westgard 4:1s, 0.472 (95% CI, 0.424–0.519) for Westgard 10x, and 0.964 (95% CI, 0.942–0.978) for the Internal QC Monitoring Agent. Median warning lead times were 2 QC results (95% CI, 2–2), or 0.037 days (95% CI, 0.028–0.044), for Westgard 1:3s or 2:2s; 6 QC results (95% CI, 5–8), or 0.481 days (95% CI, 0.281–0.651), for Westgard 4:1s; 30 QC results (95% CI, 27–33), or 4.071 days (95% CI, 3.158–5.584), for Westgard 10x; and 42 QC results (95% CI, 37–47), or 5.40 days (95% CI, 3.75–6.07), for LabARIA (eTable 1). LabARIA had the longest median warning lead time in both QC results and days among the evaluated methods.

Across the evaluated threshold settings, action recall ranged from 0.960 to 0.976, and median warning lead time ranged from 40 to 46 QC results. These findings indicate that the Internal QC Monitoring Agent's action recall and warning lead time were not sensitive to the tested monitoring thresholds (eTable 2).

##### Assay-Specific PBRTQC Monitoring

Across the 35 analytes, Bull's trimmed mean was selected for 10 analytes, rolling mean for 9, CUSUM for 9, EWMA for 4, and rolling median for 3 (eTable 3). All selected methods met the prespecified false-alert criterion, with false-alert frequencies ranging from 0 to 1.69 per 1000 patient results before the simulated shift. Under the same simulated 5% mean shift, ANPed ranged from 8.0 to 99.8 patient results and sensitivity ranged from 0.45 to 1.00 across analytes (eTable 3 and eFigure 3A). These results show that LabARIA can automatically adapt PBRTQC monitoring to individual analytes by selecting among candidate methods based on analyte-specific detection performance while meeting the prespecified false-alert criterion, which is an important capability given the observed differences in patient-result distributions and monitoring performance across analytes.

##### PBRTQC Complemented IQC

Across all 422 recorded QC actions, IQC generated warnings before 407 actions. Of the 15 actions not identified by IQC, 12 had patient-result streams available for the same analyte and instrument, and PBRTQC identified all 12. The remaining 3 actions did not have corresponding patient-result streams available for PBRTQC evaluation. Adding PBRTQC therefore increased action recall from 407 of 422 (0.964 [95% CI, 0.942–0.978]) to 419 of 422 (0.993 [95% CI, 0.979–0.998]), an absolute increase of 0.028 (95% CI, 0.014–0.045) (eTable 4 and eFigure 3B). Patient-result streams for the same analyte and instrument were available for 200 of the 422 recorded QC actions. Within these 200 actions, 182 were identified by both IQC and PBRTQC, 6 by IQC alone, and 12 by PBRTQC alone, while none were missed by both approaches (eTable 4). These findings show that PBRTQC provided complementary warning coverage for recorded QC actions not identified by IQC.

#### eResults 2. Inventory and Staffing Forecasting

Across 49 days of synthetic operational data, the Inventory Monitoring Agent tracked the number and severity of inventory items at risk and changes in days of supply, including trajectories preceding simulated stockouts and delivery delays. The Staffing Monitoring Agent tracked staffing coverage relative to required staffing across laboratory sections and shifts and identified changes in coverage over time for individual section-shift combinations (eFigure 4A-D). These findings show how the Inventory Monitoring Agent and Staffing Monitoring Agent identify changing operational risks and present them with the corresponding inventory and staffing measurements.

The Forecasting & Optimization Agent selected a forecasting method separately for each outcome and time window based on AUROC. For inventory risk forecasting, AR(1) was selected for the 3-day time window, while SMA was selected for the 7- and 14-day time windows. For staffing risk forecasting, SMA was selected for the 3- and 7-day time windows, while AR(1) was selected for the 14-day time window (eTable 5).

For inventory risk forecasting, the selected methods achieved AUROCs of 0.883 (95% CI, 0.781–0.950), 0.800 (95% CI, 0.696–0.881), and 0.719 (95% CI, 0.631–0.794) for the 3-, 7-, and 14-day time windows, respectively. For staffing risk forecasting, the corresponding AUROCs were 0.897 (95% CI, 0.792–0.969), 0.877 (95% CI, 0.778–0.950), and 0.820 (95% CI, 0.670–0.939) (eTable 5). The Forecasting & Optimization Agent applied different forecasting methods across outcomes and time windows. The selected methods distinguished records followed by inventory or staffing shortages within the specified time window from those without these outcomes.

eFigure 4E-J also reports average precision, Recall@K, Spearman rank correlation, Kendall rank correlation, and nDCG. These measures evaluate complementary aspects of risk ranking, including identification of future inventory or staffing shortages among higher-ranked records and the ordering of records by predicted risk. Together with AUROC, these results characterize the Forecasting & Optimization Agent's performance in forecasting and prioritizing inventory and staffing risks across the 3-, 7-, and 14-day time windows.

### eAppendix

#### eAppendix 1. Illustrative Scenarios of Integrated Laboratory Investigation

Two illustrative scenarios based on events encountered in laboratory practice show how LabARIA could synthesize evidence to guide investigation (**Figure 5**). In the first scenario, hemolysis-related potassium rejections in the ED increase over 2 weeks relative to its historical baseline. The Patient and Specimen module identifies the affected unit, test, and timing, while the QA Coordinator finds no corresponding change in potassium IQC or PBRTQC and reviews the acceptable prior proficiency testing performance. The Orchestrator presents these findings together, directing attention to changes in blood collection practices in ED. In the second scenario, within 1 day, variability of patient calcium results and the proportion of such results outside the reference range increase, together with out-of-range IQC measurements. The QA Coordinator integrates the PBRTQC and IQC signals to prioritize a newly emerging analytical concern. With no corresponding specimen quality abnormalities identified, the dashboard directs attention to calcium assay troubleshooting. Follow-up investigation identifies a water filtration problem, and increased reagent consumption from repeated testing prompts an inventory alert. These scenarios illustrate how coordinated findings could help laboratory personnel focus on an investigation and anticipate its operational consequences.

### eTables

#### eTable 1. IQC Action Recall and Warning Lead Time

| Method | Action recall (95% CI) | Median lead, QC results (95% CI) | Median lead, days (95% CI) |
|---|---|---|---|
| Westgard 1:3s or 2:2s | 0.976 (0.957–0.987) | 2 (2–2) | 0.037 (0.028–0.044) |
| Westgard 4:1s | 0.628 (0.581–0.673) | 6 (5–8) | 0.481 (0.281–0.651) |
| Westgard 10x | 0.472 (0.424–0.519) | 30 (27–33) | 4.071 (3.158–5.584) |
| LabARIA | 0.964 (0.942–0.978) | 42 (37–47) | 5.40 (3.75–6.07) |

Action recall was the proportion of the recorded QC actions preceded by a warning within the previous QC results. Warning lead time was measured from the onset of the earliest warning to the recorded action.

**eTable 2. Sensitivity of IQC Action Recall and Warning Lead Time to Monitoring Thresholds**

| Parameter varied | Value | Action recall | Median warning lead, QC results |
|---|---|---|---|
| Exponentially weighted moving average limit | 0.75 | 0.967 | 42 |
| Exponentially weighted moving average limit | 1.00 (default) | 0.964 | 42 |
| Exponentially weighted moving average limit | 1.25 | 0.962 | 42 |
| Cumulative sum decision interval | 3.00 | 0.967 | 42 |
| Cumulative sum decision interval | 4.00 (default) | 0.964 | 42 |
| Cumulative sum decision interval | 5.00 | 0.964 | 41 |
| Rolling standard-deviation or coefficient-of-variation limit | 1.25 | 0.976 | 46 |
| Rolling standard-deviation or coefficient-of-variation limit | 1.50 (default) | 0.964 | 42 |
| Rolling standard-deviation or coefficient-of-variation limit | 1.75 | 0.960 | 40 |

**eTable 3. Analyte-Specific PBRTQC Methods and Performance Under a 5% Mean Shift**

| Analyte | Selected method | ANPed | False alerts per 1000 results | Sensitivity |
|---|---|---|---|---|
| Calcium | Bull's trimmed mean | 50.1 | 0.21 | 0.84 |
| Sodium | EWMA | 8.0 | 0.97 | 1.00 |
| Potassium | Rolling mean | 57.0 | 0.18 | 0.82 |
| Glucose | CUSUM | 88.0 | 1.69 | 0.61 |
| Creatinine | CUSUM | 84.9 | 0.41 | 0.63 |
| Carbon dioxide | Rolling mean | 67.1 | 0.36 | 0.77 |
| Blood urea nitrogen | CUSUM | 87.7 | 0.79 | 0.59 |
| Chloride | EWMA | 16.5 | 0.82 | 0.99 |
| Albumin | Bull's trimmed mean | 78.1 | 0.54 | 0.59 |
| Total protein | EWMA | 67.8 | 0.72 | 0.70 |
| Alkaline phosphatase | Rolling mean | 85.9 | 0.38 | 0.55 |
| Total bilirubin | CUSUM | 96.1 | 0.38 | 0.52 |
| Alanine aminotransferase | Bull's trimmed mean | 82.3 | 0.87 | 0.59 |
| Aspartate aminotransferase | CUSUM | 92.1 | 0.36 | 0.56 |
| Magnesium | Bull's trimmed mean | 56.3 | 0.21 | 0.82 |
| Troponin T | CUSUM | 77.6 | 0.15 | 0.68 |
| Phosphorus | Bull's trimmed mean | 83.4 | 0.23 | 0.58 |
| Direct bilirubin | CUSUM | 91.7 | 0.44 | 0.55 |
| Lactate dehydrogenase | Rolling median | 99.8 | 0.54 | 0.45 |
| Triglycerides | CUSUM | 86.4 | 0.79 | 0.62 |
| Thyroid-stimulating hormone | Rolling median | 94.8 | 1.21 | 0.47 |
| Hemoglobin | Bull's trimmed mean | 86.5 | 0.31 | 0.58 |
| Hematocrit | Rolling mean | 83.7 | 0.18 | 0.58 |
| White blood cell count | Rolling mean | 96.9 | 0.10 | 0.50 |
| Red blood cell count | Rolling mean | 91.3 | 0.38 | 0.53 |
| Mean corpuscular volume | Bull's trimmed mean | 29.6 | 0.15 | 0.96 |
| Mean corpuscular hemoglobin | Bull's trimmed mean | 29.2 | 0.18 | 0.97 |
| Mean corpuscular hemoglobin concentration | EWMA | 19.1 | 0.49 | 0.99 |
| Red cell distribution width | Bull's trimmed mean | 77.4 | 0.38 | 0.70 |
| Platelet count | Rolling median | 91.6 | 0.79 | 0.53 |
| Mean platelet volume | Bull's trimmed mean | 47.6 | 0.15 | 0.90 |
| Total cholesterol | Rolling mean | 89.7 | 0.08 | 0.58 |
| High-density lipoprotein cholesterol | Rolling mean | 96.7 | 0.05 | 0.53 |

| Analyte | Selected method | ANPed | False alerts per 1000 results | Sensitivity |
|---|---|---|---|---|
| Low-density lipoprotein cholesterol | Rolling mean | 92.7 | 0.05 | 0.55 |
| Carcinoembryonic antigen | CUSUM | 57.2 | 0.00 | 0.80 |

For each analyte, the selected method had the lowest average number of patient results from shift onset to detection (ANPed) among methods producing no more than 3 false alerts per 1000 patient results before the simulated shift. Sensitivity was the proportion of simulated shifts detected.

**eTable 4. IQC and PBRTQC Action Recall**

| Evaluation | Measure | Result |
|---|---|---|
| All recorded QC actions (n = 422) | IQC action recall | 407/422 (0.964, 95% CI, 0.942–0.978) |
| | Combined IQC and PBRTQC action recall | 419/422 (0.993, 95% CI, 0.979–0.998) |
| | IQC misses identified by PBRTQC | 12/15 (80.0%) |
| | Absolute increase in action recall | 0.028 (95% CI, 0.014–0.045) |
| Actions with patient-result streams (n = 200) | PBRTQC action recall | 194/200 (0.970, 95% CI, 0.936–0.986) |
| | Identified by both IQC and PBRTQC | 182 |
| | Identified by IQC alone | 6 |
| | Identified by PBRTQC alone | 12 |
| | Not identified by either method | 0 |

**eTable 5. Inventory and Staffing Forecasting Performance**

| Outcome | Forecast time window, days | Selected method | AUROC (95% CI) |
|---|---|---|---|
| Inventory | 3 | First-order autoregressive model | 0.883 (0.781–0.950) |
| Inventory | 7 | Simple moving average (SMA) | 0.800 (0.696–0.881) |
| Inventory | 14 | Simple moving average (SMA) | 0.719 (0.631–0.794) |
| Staffing | 3 | Simple moving average (SMA) | 0.897 (0.792–0.969) |
| Staffing | 7 | Simple moving average (SMA) | 0.877 (0.778–0.950) |
| Staffing | 14 | First-order autoregressive model | 0.820 (0.670–0.939) |

For each outcome and time window, forecasting-method selection was based on AUROC calculated from observations whose outcome windows had closed by the final prediction day. AUROC indicates area under the receiver operating characteristic curve. CI indicates confidence interval.

**eTable 6. Analyte-Specific Hemolysis-related Rejection Cutoffs**

| Test | Hemolysis-related rejection cutoff, mg/dL |
|---|---|
| Lactate Dehydrogenase | 40 |
| Potassium | 100 |
| Direct Bilirubin | 60 |
| Aspartate aminotransferase (AST) | 100 |
| Alanine aminotransferase (ALT) | 350 |
| Unsaturated iron-binding capacity (UIBC) | 100 |
| High-sensitivity troponin | 100 |

**eTable 7. Test-Specific Turnaround-Time Targets**

| Test or profile | STAT/Routine ED target, min | Emergent target, min |
|---|---|---|
| BMP/BMPP | 60 | 45 |
| CMP/CMPP | 60 | — |
| Ammonia | 60 | — |
| Bilirubin, total, neonates | 60 | — |
| Blood gases and co-oximetry | 15 | — |
| HCG, beta, serum | 60 | — |
| Lactate | 60 | — |
| Magnesium | 60 | — |
| Osmolality, serum | 90 | — |
| CSF glucose | 60 | — |
| CSF protein | 60 | — |
| Troponin I | 60 | — |
| Urinalysis, automated | 30 | — |
| Urinalysis, manual | 60 | — |
| Urine pregnancy | 30 | — |
| CBC | 60 | 30 |
| CBCC, automated differential | 60 | — |
| CBCC, manual differential or scan | 90 | — |
| CSF cell count | 60 | — |
| Routine coagulation | 60 | 60 |
| Therapeutic drug monitoring | 120 | — |
| Ethanol | 60 | — |

TAT was measured from laboratory receipt to result verification. A TAT miss occurred when a result exceeded the applicable target. All targets are in minutes. BMP indicates basic metabolic panel. CBC indicates complete blood count. CBCC indicates complete blood count with differential. CMP indicates comprehensive metabolic panel. CSF indicates cerebrospinal fluid. ED indicates emergency department. HCG indicates human chorionic gonadotropin. TAT indicates turnaround time. A dash denotes no applicable target.

### eFigures

#### eFigure 1. LabARIA Dashboard for Laboratory Personnel

**A**, Overview of laboratory monitoring findings and priority-tiered alerts. **B**, Patient and specimen monitoring. **C**, Workload and turnaround-time (TAT) monitoring, including TAT miss rates and tests contributing to TAT misses. **D**, Chatbot-based follow-up questions about dashboard findings and supporting analytical results. **E**, Internal quality control (IQC) measurements and assay-level monitoring findings. **F**, Patient-based real-time quality control (PBRTQC) monitoring and patient-result patterns by assay. **G**, Proficiency-testing/external quality-assessment (PT/EQA) results and assay-level findings. **H**, Inventory and staffing monitoring, including available supplies, staffing coverage, and forecasting results. Displayed values illustrate the interface.

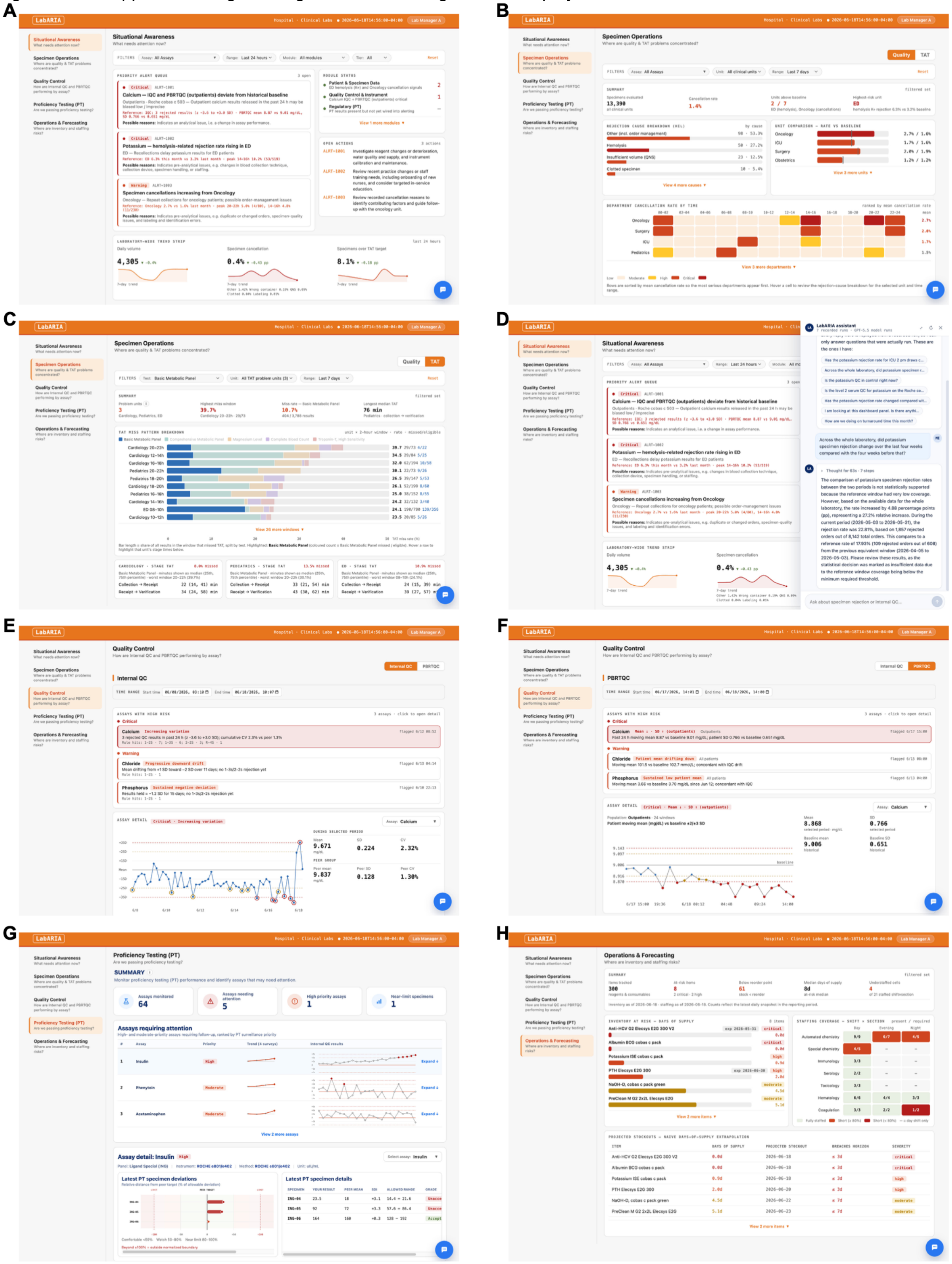

**eFigure 2. Laboratory Data Sources and Agent Architecture**

Laboratory records from laboratory results and specimens, internal quality control (IQC) reports, proficiency-testing/external quality-assessment (PT/EQA) reports, maintenance and operational logs, and laboratory rules are prepared by routing agents. Foundation agents provide analytical and reasoning capabilities used by the application agents. Application agents perform laboratory monitoring and integrate analytical findings within the three functional modules. The Orchestrator coordinates analytical tasks and assembles findings with supporting measurements and recommended checks for laboratory personnel. IQC indicates internal quality control. PBRTQC indicates patient-based real-time quality control. PT/EQA indicates proficiency testing/external quality assessment. QC indicates quality control. TAT indicates turnaround time.

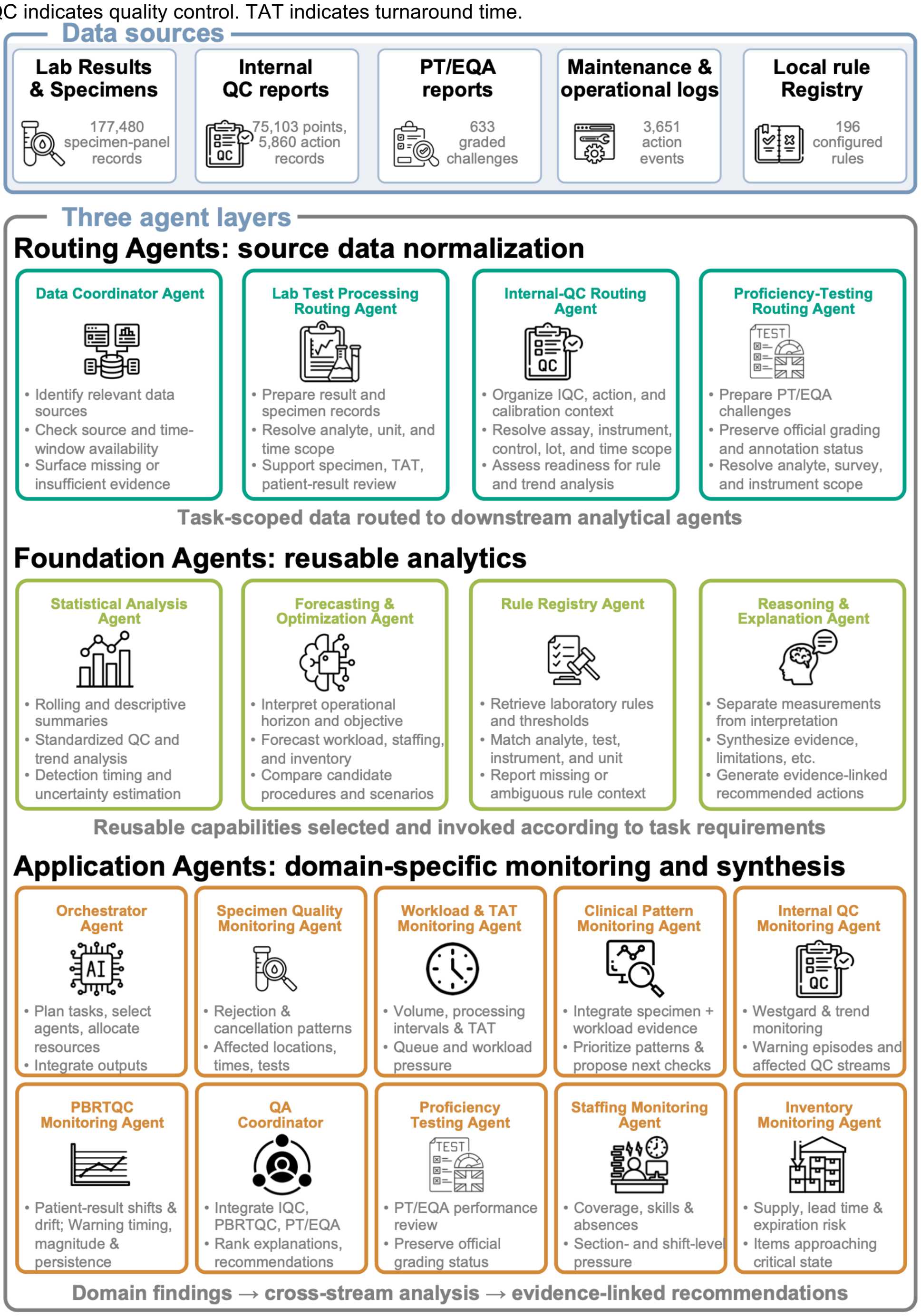

#### eFigure 3. PBRTQC Monitoring Performance and Complementary Warnings

**A**, PBRTQC monitoring performance across 35 analytes after a simulated 5% mean shift. Points indicate false-alert frequency before the shift and average number of patient results from shift onset to detection (ANPed) for the method selected for each analyte. Lines connect points to their analyte labels. Colors indicate rolling mean, rolling median, exponentially weighted moving average (EWMA), cumulative sum (CUSUM), and Bull's trimmed mean. Each method was selected to minimize ANPed while producing no more than 3 false alerts per 1000 patient results before the shift. **B**, Action recall for IQC alone and combined IQC and PBRTQC monitoring across 422 recorded QC actions. IQC identified 407 actions (recall, 0.964), and combined monitoring identified 419 (recall, 0.993), including 12 additional actions identified by PBRTQC. Error bars indicate 95% confidence intervals (CIs). CUSUM indicates cumulative sum. EWMA indicates exponentially weighted moving average. IQC indicates internal quality control. PBRTQC indicates patient-based real-time quality control. QC indicates quality control.

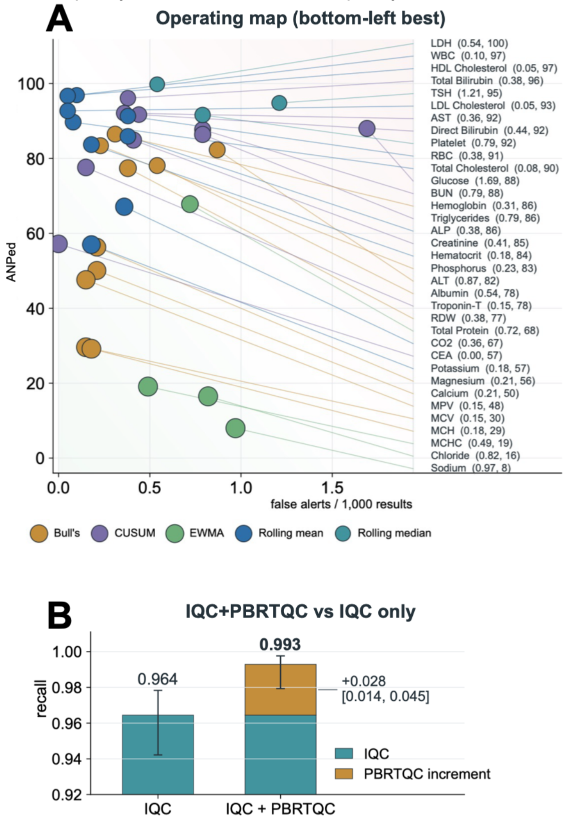

#### eFigure 4. Inventory and Staffing Monitoring and Forecasting

**A-D**, Inventory and staffing monitoring using 49 days of synthetic operational data. **A**, Numbers of inventory items at risk by day and severity. **B**, Days of supply for creatinine and potassium, illustrating simulated stockouts and delivery delays. **C**, Staffing coverage across laboratory sections and shifts. **D**, Staffing coverage over time for a selected laboratory section and shift, compared with the required level. **E-G**, Inventory forecasting performance at 3 (**E**), 7 (**F**), and 14 (**G**) days. **H-J**, Staffing forecasting performance at 3 (**H**), 7 (**I**), and 14 (**J**) days. Each forecasting panel compares the forecasting method selected by the Forecasting & Optimization Agent with random scores (denoted as random in the figure), time since restocking or adequate staffing (denoted as overdue), and historical event frequency (denoted as frequency).

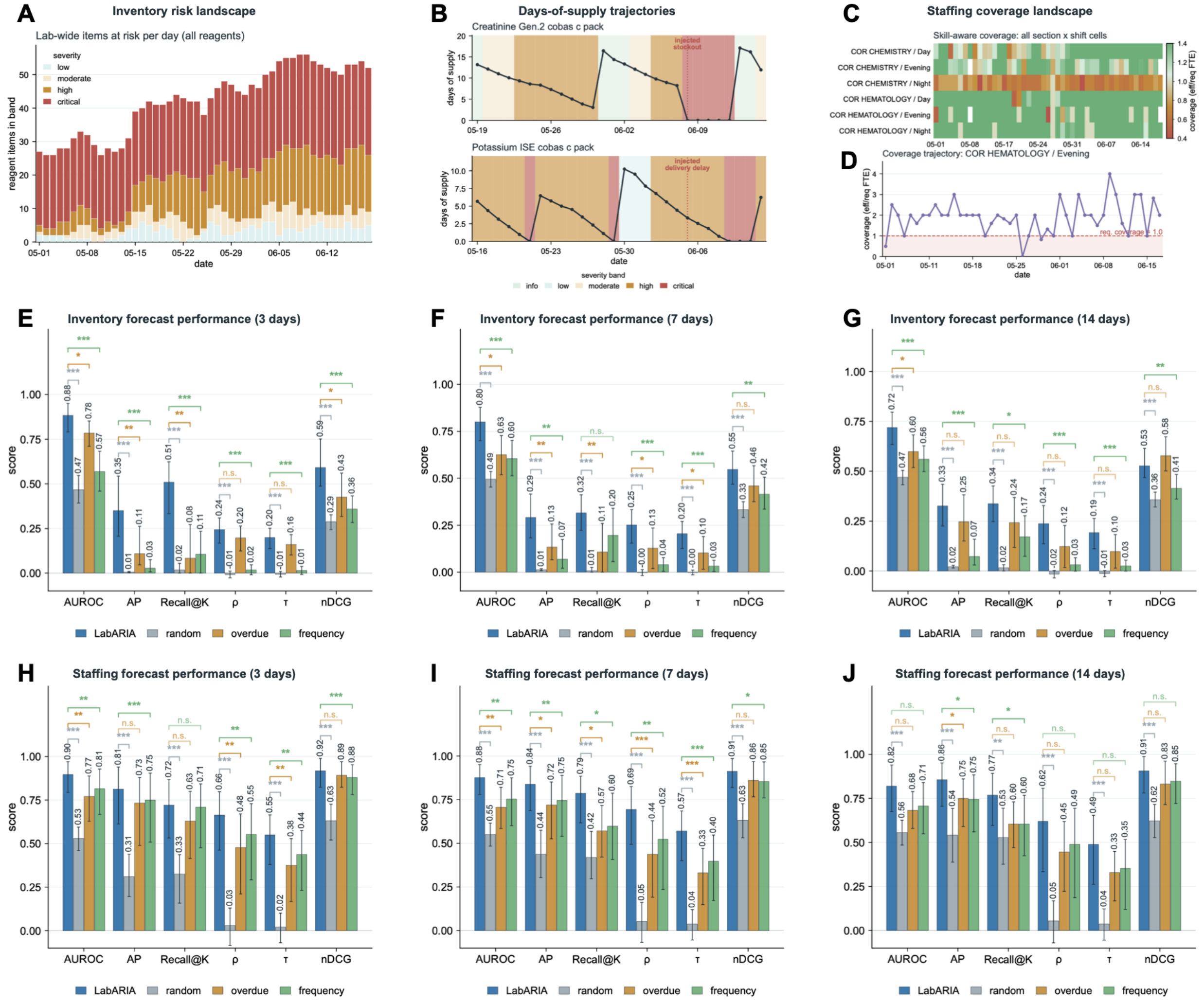
